# Multiple-Input Deep Convolutional Neural Network Model for COVID-19 Forecasting in China

**DOI:** 10.1101/2020.03.23.20041608

**Authors:** Chiou-Jye Huang, Yung-Hsiang Chen, Yuxuan Ma, Ping-Huan Kuo

## Abstract

COVID-19 is spreading all across the globe. Up until March 23, 2020, the confirmed cases in 173 countries and regions of the globe had surpassed 346,000, and more than 14,700 deaths had resulted. The confirmed cases outside of China had also reached over 81,000, with over 3,200 deaths. In this study, a Convolutional Neural Network (CNN) was proposed to analyze and predict the number of confirmed cases. Several cities with the most confirmed cases in China were the focus of this study, and a COVID-19 forecasting model, based on the CNN deep neural network method, was proposed. To compare the overall efficacies of different algorithms, the indicators of mean absolute error and root mean square error were applied in the experiment of this study. The experiment results indicated that compared with other deep learning methods, the CNN model proposed in this study has the greatest prediction efficacy. The feasibility and practicality of the model in predicting the cumulative number of COVID-19 confirmed cases were also verified in this study.

## 1. Introduction

In early December 2019, the first case of COVID-19 infection was discovered in Wuhan City in China’s Hubei Province [1]. In the following weeks, this disease broke out within China and continued to spread extensively in other countries, causing worldwide panic. The pneumonia caused by this novel coronavirus was officially named the Coronavirus disease 2019 (COVID-19) by the World Health Organization (WHO). The 2019 coronavirus epidemic was triggered by the Severe Acute Respiratory Syndrome Coronavirus 2 (SARS-CoV-2), and it has, at the time of writing, spread in over 173 countries, causing more than 15,000 deaths. The epidemic is still spreading, and the WHO has classified the global risk level of the disease as a “pandemic.” Currently, the earliest known case symptoms appeared on December 1, 2019, and the first case sought treatment on December 8. On December 26, 2019, Wuhan City Respiratory and Intensive care doctor Zhang Jixian first discovered this pneumonia with unknown cause and suspected that it is an infectious disease. Subsequently, the disease broke out in Wuhan City. On January 20, 2020, Chinese academic Zhong Nanshan publicly announced that the novel coronavirus pneumonia “definitely transmits between people.” On January 23, 2020, the Wuhan City Government announced the adoption of lockdown and quarantine measures in the infected areas, which was the first case of lockdown in a major city (with a population of 11 million) in recent public health history. Since January 13, the disease continued to spread to Thailand, Japan, and South Korea. On January 30, three countries outside China were verified as having interpersonal propagation, and the pandemic was thus designated as an international public health emergency event by the WHO.

According to reports on the epidemic, “the propagation speed is faster, and the virus propagation power has increased.” Infected people can infect others with the virus while exhibiting no symptoms; and the latency from infection to the presentation of symptoms is as long as 14 days. These characteristics also increase the difficulty of controlling the epidemic. As the epidemic continues, there is also a problem of the global undersupply of surgical masks. Currently, no vaccine and remedy for the novel coronavirus have been discovered. WHO assistant director-general Bruce Aylward stated that Remdesivir is currently the only drug that is “considered to probably have real efficacy.” The China–Japan Friendship Hospital in China and the American National Institute of Allergy and Infectious Diseases are starting to conduct clinical trials for the drug. At present, no sufficient knowledge regarding the disease is known, and key factors, such as virus source, virus birthplace, morbid mechanism, virus pathogenicity, and propagation power are still uncertain. The WHO stated on March 3 that with the novel coronavirus epidemic, the world is in an “unknown state.”

Up until March 23, 2020, 345,297 people have been infected with the virus; among these people, 14,765 have died. The several European countries with the most severe epidemics have experienced the maximum increase in confirmed cases in a single day on March 5: cases for Italy increased from 3,089 to 3,858, those for Germany increased from 262 to 482, those for France increased from 285 to 423, and those for the Netherlands increased more than twofold from 38 to 82. In the WHO report on February 26, the numbers of new cases in China and Japan have decreased, but the numbers of new cases in Italy, Iran, and South Korea are still rising. At the time of writing, the numbers of confirmed cases in Germany, France, and Spain have surpassed 1,000.

## 2. Related Works

Fig. 1 [2] presents the global map of COVID-19 occurrence according to data from the United States’ Centers for Disease Control and Prevention. The countries marked with a color are countries with confirmed cases. At present, 173 of the 195 countries in the world have confirmed cases, underscoring the epidemic’s astounding speed of propagation from its origin in China’s Hubei province. Most scholars have used mathematical models to predict the spread of COVID-19. Roosa et al. [3] used the generalized logistic model, the Richards model, and the sub-epidemic model to establish prediction models for the cumulative numbers of confirmed cases in Guangdong Province and Zhejiang Province. They predicted that, in the next 5 days and 10 days from their time of writing, 65–81 and 44–354 new cases will appear in Guangdong and Zhejiang provinces, respectively. In addition, Liu et al. [4] used mathematical models to simulate the propagation status of COVID-19 in China and noted the importance of the government’s policy of limiting public activities and the movement of infected people with no symptoms. Boldog et al. [5] adopted a Time-Dependent Compartmental Model of the Transmission Dynamics to estimate the cumulative number of confirmed cases outside of Hubei Province. Moreover, the Galton–Watson branch was used to analogize virus propagation, thus determining the connectivity and transmissibility between China and the destination country. The disadvantage of this method is that the uncertainty regarding the efficacy of control measures against the disease results in difficulties in predicting the development trajectories of the disease. By comparison, Al-Qaness et al. [6] adopted the adaptive neuro-fuzzy inference system (ANFIS), an improvement from flower pollination algorithm and salp swarm algorithm, to predict the number of confirmed cases in the next 10 days. The comparison with the ANFIS optimized with GA, PSO, ABC, and FPA indicated that the accuracy of the FPASSA-ANFIS model reached 0.97, and the predicted average increase in numbers of new cases in the next 10 days was 10% more than the number of present cases. Jung et al. [7] used the rate of increase together with delayed distribution estimation and statistical induction to establish a mathematical model, based on COVID-2019 case data reported before January 24. The predicted cumulative number of confirmed cases for up until January 24 was 6,924, and the predicted death ratio was 5.3%; these figures were within the 95% confidence interval. Compared with previous methods for establishing mathematical models, Fan et al. [8] adopted a statistical method for predicting the floating population in Wuhan City. The experiment indicated that the floating population in Wuhan region is highly correlated with the number of daily confirmed cases. The residence time of the floating population for local cases was longer than that for non-local cases, which results in a lower predicted number of confirmed cases in the areas around Hubei Province. The prediction results indicate that approximately 80% of the epidemic will be centralized in the top 30 districts.

**Fig. 1.**
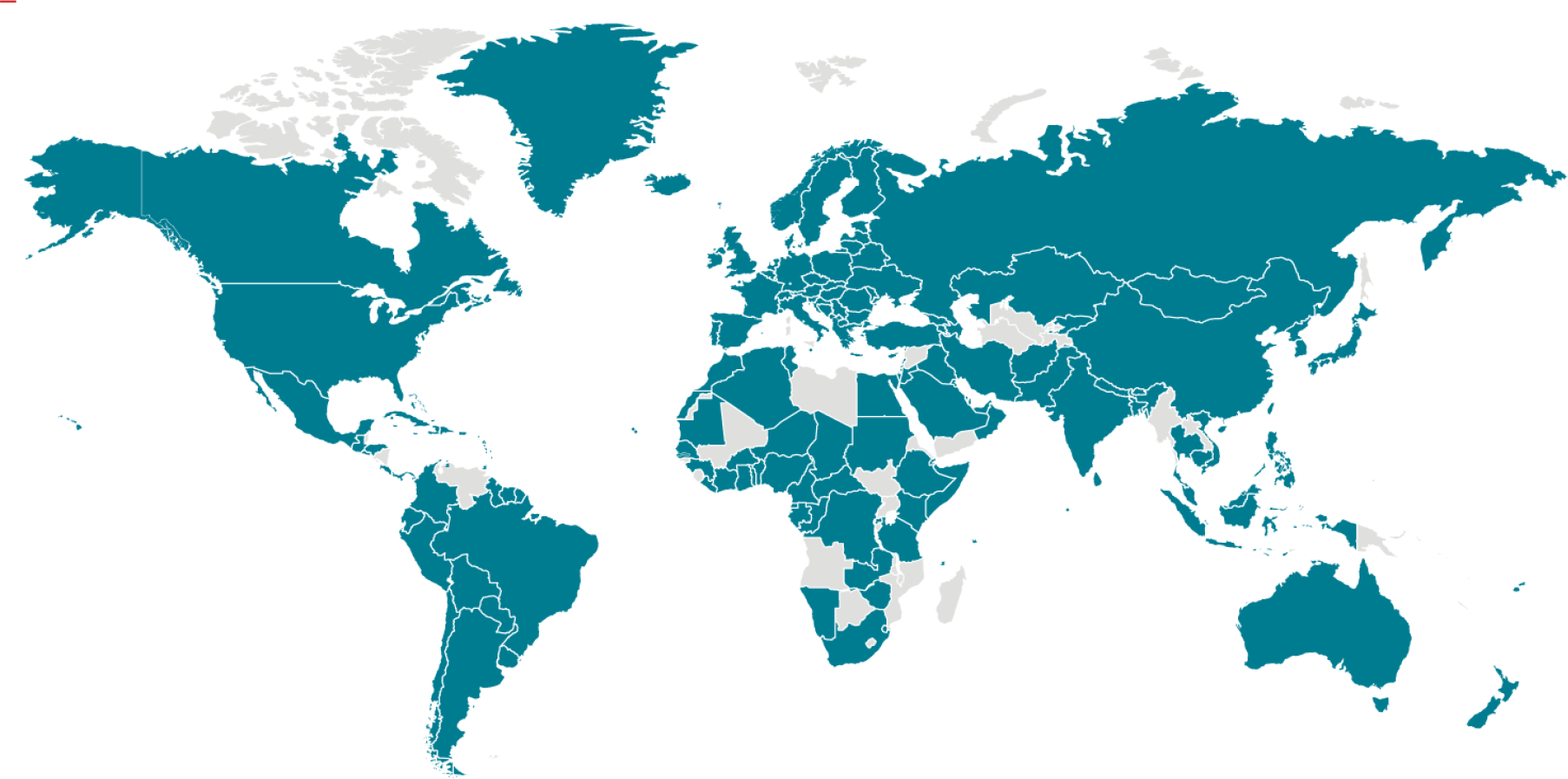
Global Map of Confirmed COVID-19 Cases as of 12:00 a.m. ET, March 21, 2020 [2].

However, applications of deep learning methods have mainly used the genome of the 2019-cCoV virus to predict disease propagation. Yang et al. [9] used the Susceptible-Exposed-Infectious-Removed model to integrate the epidemic curve, combining it with the long short-term memory model. The maximum value was reached on February 28, and the curve gradually lowered in late April. Hu et al. [10] used an improved stacked autoencoder and a cluster algorithm to group the instantaneous confirmed cases in every province; they noted a high accuracy in AI-based methods for COVID-19 trajectory prediction. The epidemic was predicted to end in mid-April. Guo et al. [11] used deep learning–based virus host prediction to compare the gene sequence of 2019-nCoVs with those of Severe Acute Respiratory Syndrome Coronavirus (SARS-CoV), bat SARS corona virus, and Middle East respiratory syndrome–related coronavirus (MERS-CoV). The bat SARS coronavirus was discovered to have a more similar mode of infection to COVID-19. Metsky et al. [12] used a nucleic acid test based on clustered regularly interspaced short palindromic repeats (CRISPR) genome editing and selected 67 viruses and subspecies as well as subspecies that are highly similar to 2019-nCoV. Thus, 67 test methods were designed in the ADAPT system to increase the sensitivity and speed of virus detection.

Riou et al. [13] simulated the early outbreak trajectories of 2019-nCoV and noted that the basic reproduction number *R* of 2019-nCoV was approximately 2.2 (90% high density interval 1.4–3.8) and that 2019-nCoV has the potential for continual infection between people. Liu et al. [14] compared 2019-nCoV with SARS and estimated the doubling time of 2019-nCoV and SARS as well as the basic reproduction number *r0* and time-varying instantaneous reproduction number *rt* through data analysis. They discovered that the infectivity of 2019-nCoV may be stronger than SARS, but that disease control efforts are effective. Ming et al.

[15] adopted an improved version of the Susceptible-Infectious-Recovered (SIR) model to predict the actual number of infection cases of 2019-nCoV as well as the actual load on the intensive care units under different diagnosis rates and public health intervention efficacies. They discovered that under a 50% diagnosis rate and no public health intervention, the actual number of cases will be significantly higher than the reported number, whereas under a 70% public health intervention, the load on the health system will decrease substantially. Zhao et al. [16] used a probability prediction model to accurately predict the infection nodes from snapshots of spreading. Fountain-Jones et al. [17] used machine learning to establish pathogen-risk models and compared the prediction results of different machine learning methods and explained the results by using game theory.

Benvenuto et al. [18] adopted an autoregressive integrated moving average model to predict the epidemic trend and morbidity of 2019-nCoV with a 95% confidence interval. They discovered that if the virus does not mutate, the case number will reach a plateau. Li et al. [19] used a function to separately describe the daily infection and death data of 2019-nCoV in Hubei and outside Hubei. The author thought that the inflection point in the Hubei region was at February 6, 2020, and that the epidemic will end on March 10, 2020, with an estimated infection rate of 39,000 people. However, the data did not include the data after February 12, where such inclusion will increase the predicted figures by 1.4 times. Yang et al. [9] described the clinical characteristics and imaging manifestation of the infected cases of COVID-19 that had been confirmed in the hospitals in Wenzhou area. In addition, the patients with and without travel or residence history in Hubei were compared. Tian et al. [20] analyzed the clinical and epidemiological characteristics of COVID-19 in the Beijing area. The characteristics of severe confirmed cases and common confirmed cases were compared, and the characteristics of COVID-19 and SARS were compared as well. Chen et al. [21] analyzed some cases of pregnant women with COVID-19 and noted no evidence that COVID-19 causes intrauterine infection through vertical transmission among women in late pregnancy. The major contribution of that study is its fast establishment of a prediction model by using deep learning with a small dataset, making it an important reference for other countries in their containment of the COVID-19 epidemic.

The structure of this study proceeds as follows. The first section introduces the entire study. The second section is a literature review. The following third section details this study’s proposed deep neural network algorithm. The fourth section presents and discusses the experiment results in detail. Finally, the fifth section concludes the entire study and underscores the contribution of this study and the predicted trend of the COVID-19 epidemic.

## 3. Artificial neural networks: A background

An Artificial Neural Network (ANN) [22, 23] is a mathematical model that mimics how neurons operate in organisms. It is a very powerful tool for establishing nonlinear models. An early ANN structure is the Multilayer Perceptron (MLP), which uses the neural network of fully connected structures; it has decent efficacy and numerous applications. However, if the data is too complex, simply using the MLP structure may cause the model to be unable to effectively learn how to handle every situation. At present, ANN has already developed into various new structures. The main structure of this study is the Convolutional Neural Network (CNN) [24, 25], and three other neural network structures are compared with it. These four neural network structures are detailed in the following subsections.

### 3.1. Convolutional neural network

The 1D convolution operation [26] is illustrated in Fig. 2. The difference between CNN and MLP is that CNN uses the concept of weight sharing. As illustrated in Fig. 2, the advantage of CNN is that its weight number does not have to be as large as that for a fully connected structure. Consequently, the training is relatively easier, and important characteristics are extracted more effectively. CNN is a feedforward neural network, which is an improvement from MLP. CNN contains four levels in structure: an input layer, convolutional layer, pooling layer, and fully connected layer.

**Fig. 2.**
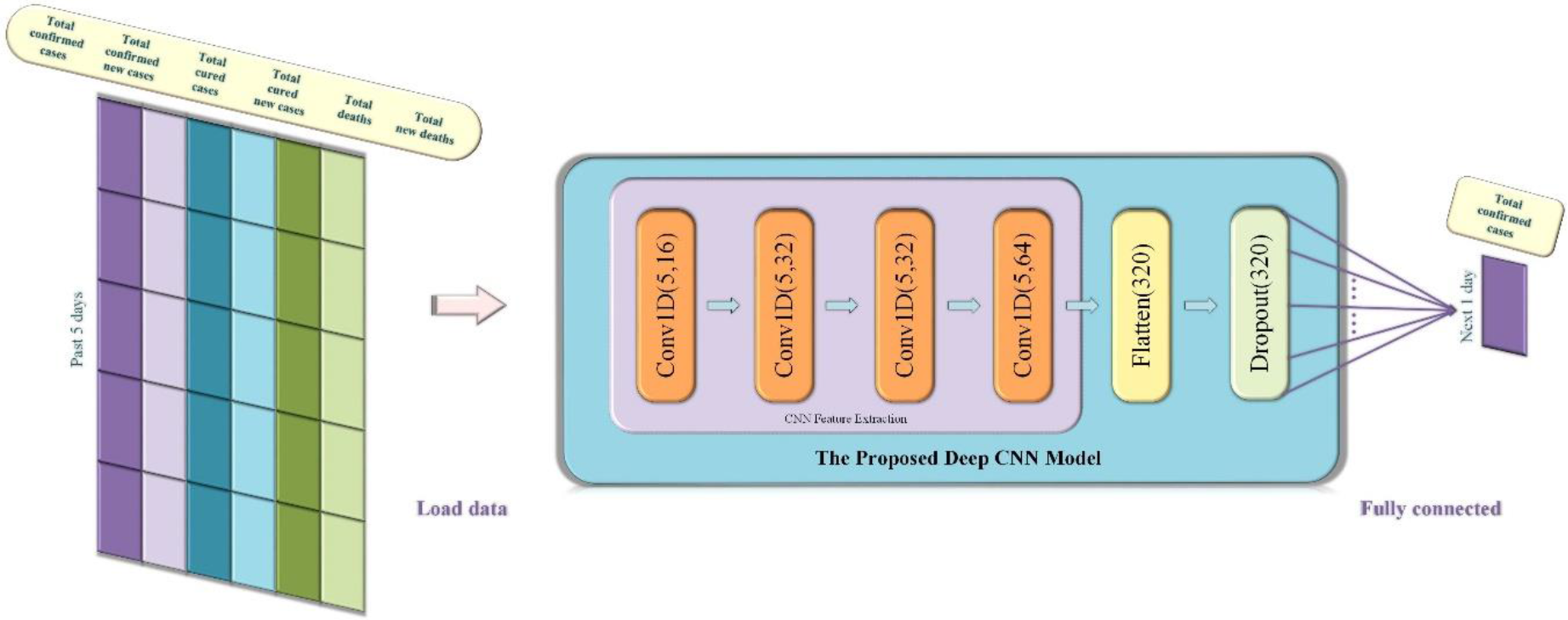
Structure of the proposed deep CNN model.

The deep CNN structure proposed in this study is illustrated in Fig. 2, and the structure is composed of CNN and the dropout layer. The input layer structure is constituted by six time sequences composed of factors influencing the cumulative number of cases. The convolutional layer structure is divided into four layers, and each convolution layer has 16, 32, 32, and 64 convolutional kernels. Going through every convolutional layer, the convolutional kernels of the previous layer slide in the input data matrixes. The convolution process can be expressed in Equation (1), in which ***X***_*ij*_ is the *i*th and *j*th matrix in the row and column direction, respectively, corresponding to the convolution kernels in the input matrix through smooth shifting. *K* is the convolution kernel, and ***γ*** is the output matrix.

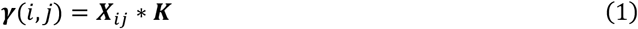

The structure principle of the fully connected layer can be expressed by Equation (2):

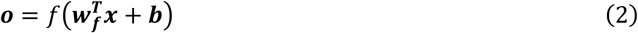

In (2), ***o*** is the vector composed of output values, ***x*** is the vector composed of input values, 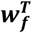 is the vector composed of weight values, ***b*** is the vector composed of threshold values, and *f* is the activation function. The output of the convolution layer can yield a 1D vector through Flatten expansion. Subsequently, the vectors are connected to the fully connected layer (dense) to obtain a 1D output. At the time of writing, the COVID-19 epidemic has continued for only 3 months, 1 week, and 3 days. Thus, considering the small number of obtained samples, this model prevents overfitting by introducing the method of dropout regularization.

### 3.2. Multilayer perceptron

A MLP is an ANN model that includes the input layer, hidden layer, and output layer. The structure of the ANN is illustrated in Fig. 3. Different connection layers are all fully connected, and the principle is similar to that of MLP. Each neural unit is connected with a weight coefficient (*w*_*i*_), and the combination of transmission result *w*_*i*_*x*_*i*_ and the deviation *b*_*i*_ is calculated using Equation (3). In addition, current neural unit output results are obtained through the activation function *f*.

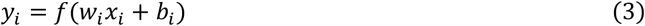

**Fig. 3.**
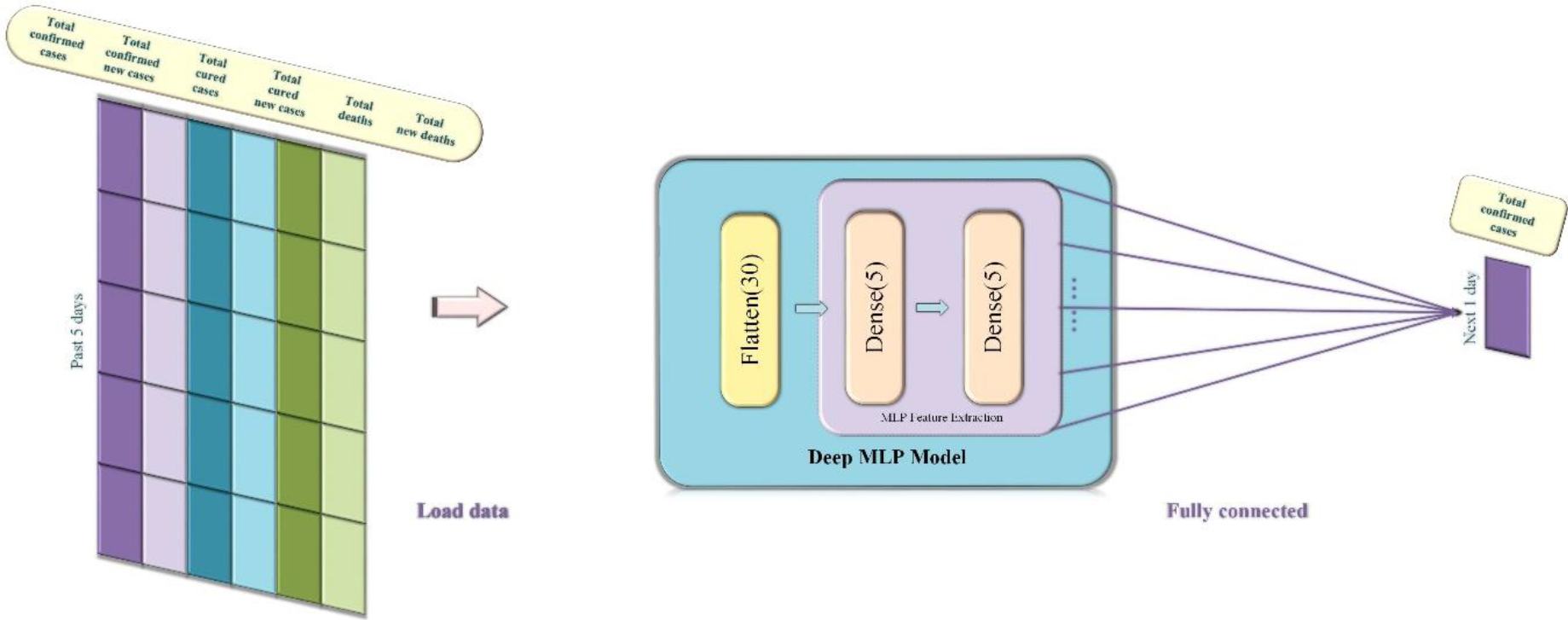
Structure of the comparative deep neural network MLP adopted in this study.

The structure of the proposed deep MLP is illustrated in Fig. 3. The structure is composed of two levels of MLP, and each MLP layer has 16 neural units. The input structure is constituted by six factors influencing the cumulative number of cases, with the time step of 5. Through a Flatten expansion, a 1 × 30 vector is derived, and the output is obtained after connecting the vector to the fully connected layer (dense).

### 3.3. Long short-term memory neural network

The long short-term memory model (LSTM) [27, 28] is an improvement from the recurrent neural network. The difference between the RNN and LSTM is that for LSTM, a cell state is added to store long-term states. In the neural unit model structure in Fig. 4 (a), the internal structure of LSTM can be divided into the input gate, forget gate, and output gate. The principle of the LSTM input gate is expressed in the following formulae. Equation (4) is used to decide which piece of information is to be added by passing *h*_*t*−1_ and *x*_*t*_ through the sigmoid layer. Subsequently, Equation (5) is used to pass *h*_*t*−1_ and *x*_*t*_ through the tanh layer to obtain new information 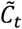. Equation (6) is used to combine the information of the current moment 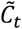 and long-term memory *C*_*t*−1_ into a new memory state *C*_*t*_.

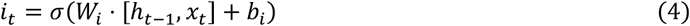

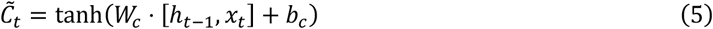

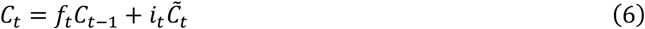

**Fig. 4.**
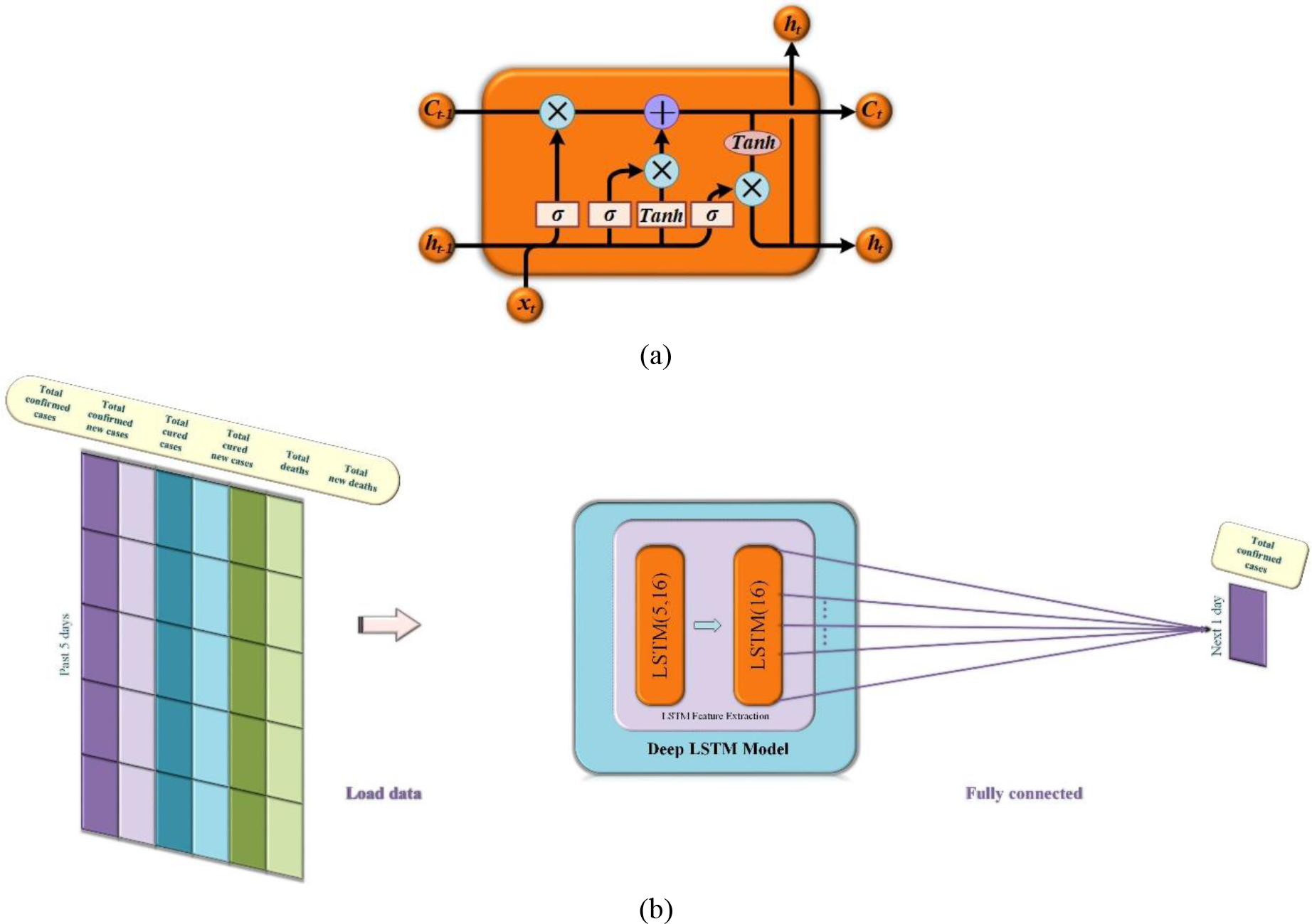
Structure of the comparative deep neural network LSTM adopted in this study. (a) LSTM neural unit; (b) the deep neural network LSTM model with an input layer consisting six input signals.

The forget gate of the LSTM uses a sigmoid layer and a dot product to allow information to pass through selectively. Equation (7) allows the LSTM to decide whether to forget the related information of the previous cell, at a certain probability, in which *W*_*f*_ is the weight matrix, and *b*_*f*_is the offset term.

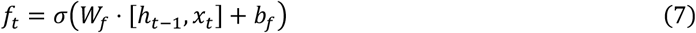

The output gate of LSTM decides which states are required to be maintained by the input *h*_*t*−1_ and *x*_*t*_ according to Equations (8) and (9). The final output results are obtained by passing the new information *C*_*t*_ through the tanh layer to multiply with state judgement vectors.

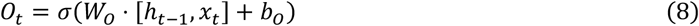

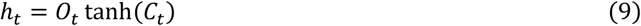

The deep LSTM structure proposed in this study is illustrated in Fig. 4 (b). The structure is composed of two LSTM layers, and each layer has 16 neural units. The input structure is constituted by six factors influencing the cumulative number of cases, with a time step of 5. A 1D output is obtained through the fully connected layer.

### 3.4. Gate recurrent unit

The Gate Recurrent Unit (GRU) [29, 30] is a variation of the LSTM, as illustrated in Fig. 5 (a). GRU changes the input gate and forget gate of LSTM into the update gate. GRU retains the effect of LSTM and simplifies the internal structure of GRU model units. The principle of GRU’s update gate is described in Equation (10). Specifically, the neural unit information of the previous moment *h*_*t*−1_ and the input data of the current moment *x*_*t*_ are combined and input to the sigmoid layer to obtain the update information of the current moment *z*_*t*_. *W* is the weight matrix.

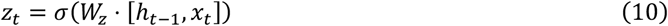

**Fig. 5.**
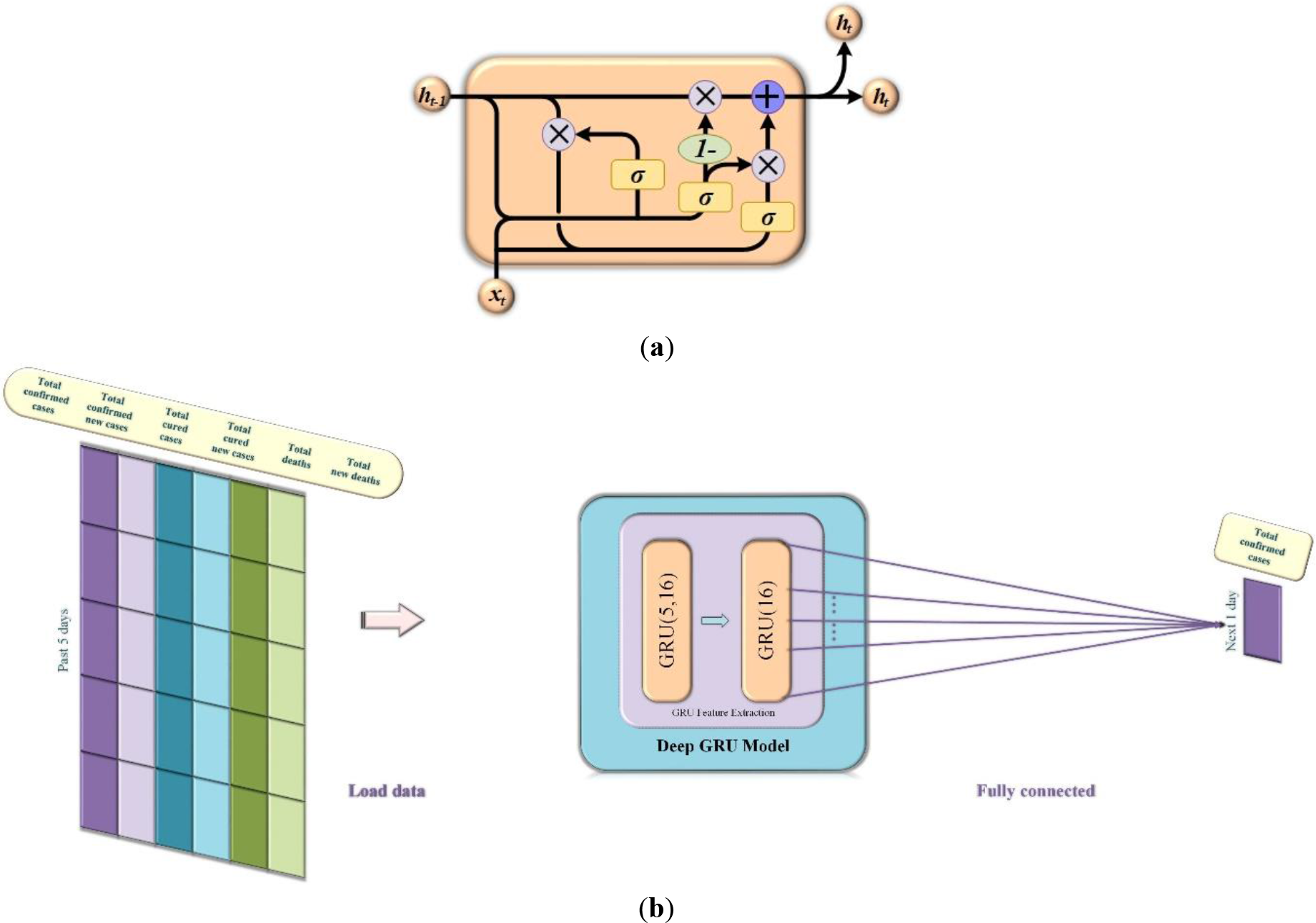
Structure of the comparative deep neural network GRU adopted in this study. (a) GRU neural unit; (b) the deep neural network GRU model with an input layer consisting six input signals.

The principle of the GRU reset gate is described in Equation (11). The reset information *r*_*t*_ is reserved after *h*_*t*−1_ and *x*_*t*_ are separately combined with the weight matrix.

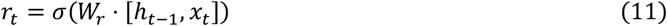

The input information of the current moment *x*_*t*_ is maintained with the obtained *r*_*t*_ at the weight probability. The following Formula (12) is used to calculate the information of the current moment 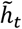.

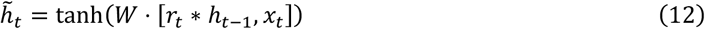

Equation (13) combines the reserved long-term memory *z*_*t*_ and the information of the current moment 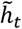 to obtain new information *h*_*t*_.

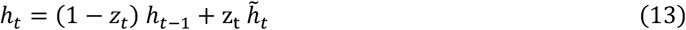

The structure of the proposed deep GRU is shown in Fig. 5 (b). The structure is composed of two GRU layers, and each layer has 16 neural units. The input structure is constituted by six factors influencing the cumulative number of cases, with a time step of 5. A 1D output is obtained through the fully connected layer.

The pseudocode of the proposed CNN is described in Algorithm 1. Specifically, the dataset and test set are first loaded. The input data are then reconstructed into 3D matrixes, and the output data format is set to the 2D matrix. The CNN model is constructed, and related CNN parameters are set. Before the training starts, the weighting value *w* is initialized, and the repeat operator *a* is initialized as 0. In the training process, the training samples are trained in batches, and the loss function is calculated once per batch to update the weight value *w*. After the training, the test set data are registered to the trained network. The errors are calculated, and the actual values and error values are compared.

#### Algorithm 1 Convolutional Neural Network

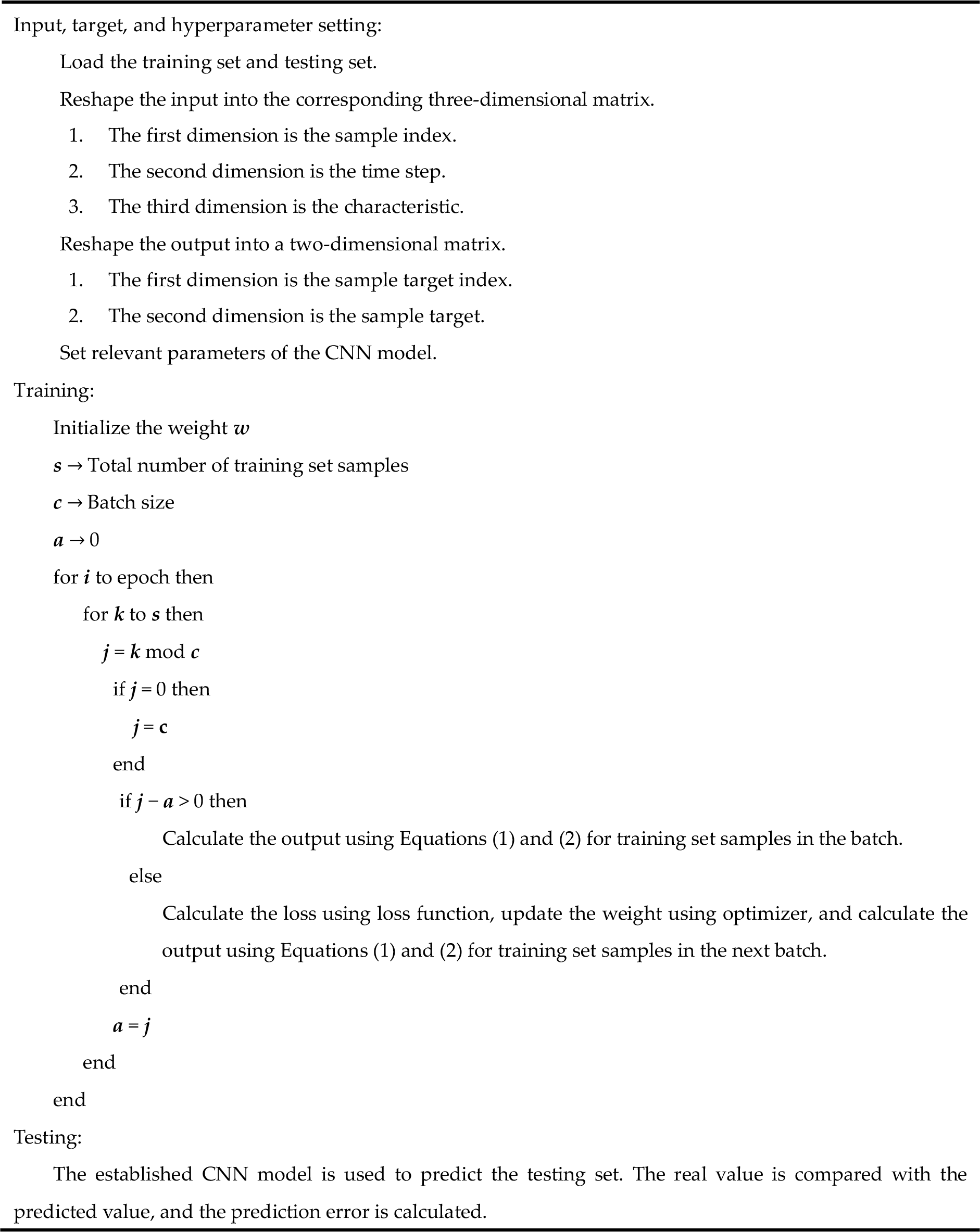

## 4. Experimental Results and Discussion

Data on confirmed cases of COVID-19 from January 23, 2020 to March 2, 2020, and from January 23, 2020 to March 2, 2020, were obtained from Surging News Network (a media outlet) [9] and WHO [31], respectively. In this experiment, the two evaluation indexes of the mean absolute error (MAE) and root mean square error (RMSE) were used; their formulae are presented in Equations (14) and (15). To conduct comprehensive efficacy tests, the dataset for January 23 to February 17 was chosen as the training data, and the dataset for February 18 to March 2 was used as testing data. Figs. 6 to 12 present the comparative prediction results of all the algorithms. The detailed MAE and RMSE values are listed in Tables I, II, III, and IV. The experiment results indicated that the GRU has decent efficacy, and that CNN is the best performing algorithm among its counterparts tested. The experiment demonstrated that the characteristic extraction of CNN is very helpful for predicting the number of confirmed cases of COVID-19.

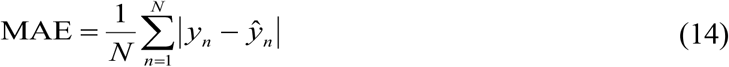

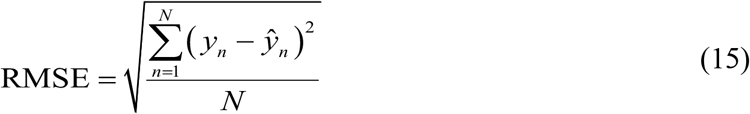

**TABLE I.** Experimental results in terms of mean absolute error of the input layer of six neurons.

| City | GRU | LSTM | MLP | CNN |
| --- | --- | --- | --- | --- |
| **Wuhan** | 17119.101 | 19671.849 | 35797.372 | **426.179** |
| **Huanggang** | 243.399 | 250.142 | 1159.923 | **89.026** |
| **Xiaogan** | 489.004 | 592.220 | 1616.017 | **87.343** |
| **Ezhou** | 259.843 | 350.798 | 814.801 | **77.942** |
| **Yichang** | 50.219 | 53.632 | 346.853 | **31.688** |
| **Wenzhou** | 15.067 | 13.988 | 120.844 | **4.617** |
| **Shenzhen** | 17.414 | 18.200 | 116.240 | **3.808** |
| **Avg.** | 2599.150 | 2992.976 | 5710.293 | **102.943** |

**TABLE II.** Experimental results in terms of mean absolute error of the input layer of a single neuron.

| City | GRU | LSTM | MLP | CNN |
| --- | --- | --- | --- | --- |
| **Wuhan** | 12054.218 | 21669.078 | 36307.670 | **1131.627** |
| **Huanggang** | 450.634 | 405.468 | 1206.190 | **255.561** |
| **Xiaogan** | 580.525 | 717.547 | 1629.771 | **389.392** |
| **Ezhou** | 168.118 | 331.491 | 887.933 | **102.507** |
| **Yichang** | 109.966 | 96.664 | 376.636 | **63.651** |
| **Wenzhou** | **20.417** | 22.303 | 141.863 | 22.138 |
| **Shenzhen** | 34.452 | 29.589 | 129.942 | **27.136** |
| **Avg.** | 1916.904 | 3324.591 | 5811.429 | **284.573** |

**TABLE III.** Experimental results in terms of root mean square error of the input layer of six neurons.

| City | GRU | LSTM | MLP | CNN |
| --- | --- | --- | --- | --- |
| Wuhan | 17140.879 | 19681.133 | 35829.022 | **456.910** |
| Huanggang | 243.715 | 250.594 | 1160.173 | **94.271** |
| Xiaogan | 490.381 | 593.989 | 1617.454 | **90.685** |
| Ezhou | 260.019 | 351.126 | 815.025 | **81.469** |
| Yichang | 50.960 | 54.844 | 347.112 | **32.855** |
| Wenzhou | 15.264 | 14.051 | 120.845 | **5.106** |
| Shenzhen | 17.446 | 18.217 | 116.241 | **4.779** |
| Avg. | 2602.666 | 2994.851 | 5715.125 | **109.439** |

**TABLE IV.** Experimental results in terms of root mean square error of the input layer of a single neuron.

| City | GRU | LSTM | MLP | CNN |
| --- | --- | --- | --- | --- |
| Wuhan | 12101.919 | 21715.728 | 36338.153 | **1416.082** |
| Huanggang | 451.136 | 406.029 | 1206.424 | **256.246** |
| Xiaogan | 583.509 | 720.488 | 1631.175 | **391.768** |
| Ezhou | 168.461 | 331.896 | 888.127 | **102.974** |
| Yichang | 110.629 | 97.439 | 376.867 | **64.649** |
| Wenzhou | 20.417 | 22.303 | 141.863 | **22.139** |
| Shenzhen | 34.454 | 29.591 | 129.943 | **27.139** |
| Avg. | 1924.361 | 3331.925 | 5816.079 | **325.857** |

**Fig. 6.**
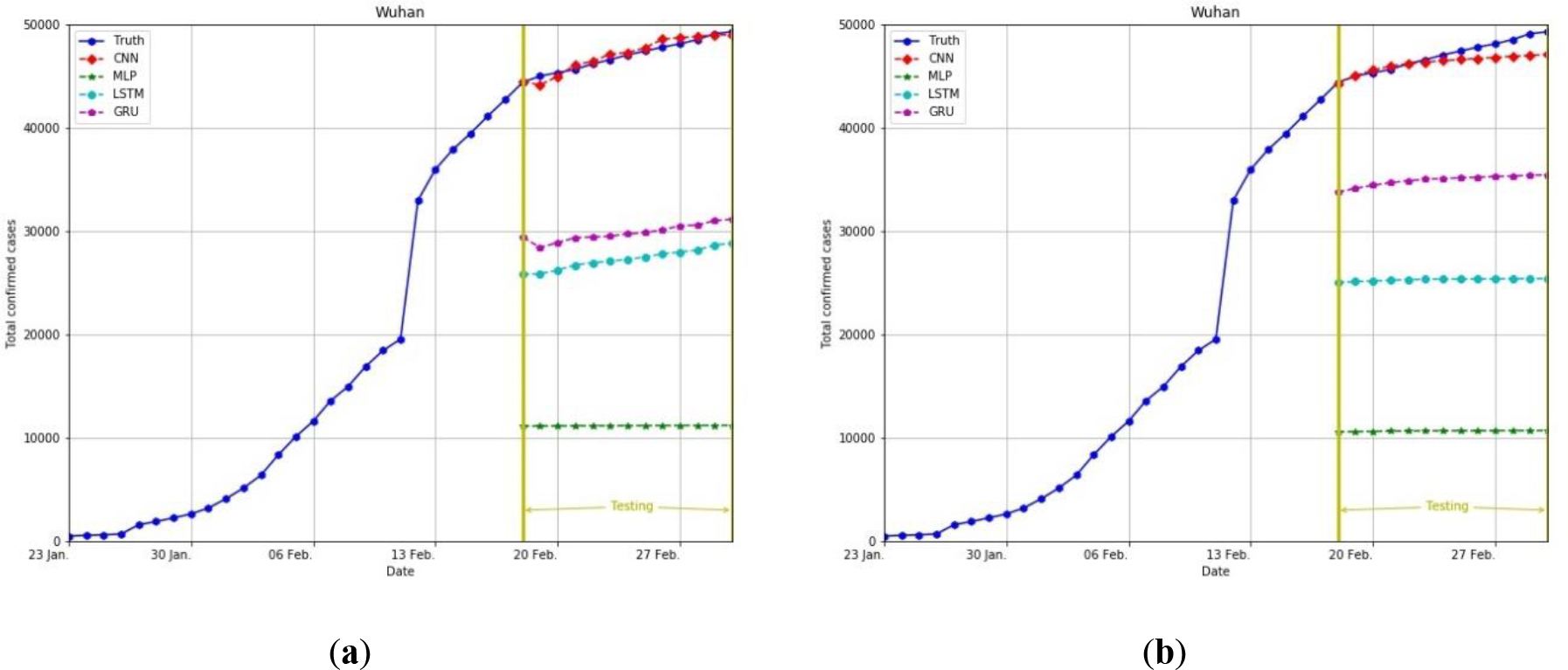
Comparison of prediction results for the cumulative number of confirmed cases in Wuhan City, Hubei Province, China. The comparison used the deep neural networks CNN, MLP, LSTM, and GRU. (a) Six important factors were adopted as the input layer: new confirmed cases, new deceased cases, new cured cases, cumulative confirmed cases, cumulative deceased cases, and cumulative cured cases. (b) Only one important factor was adopted as the input layer: cumulative confirmed cases.

**Fig. 7.**
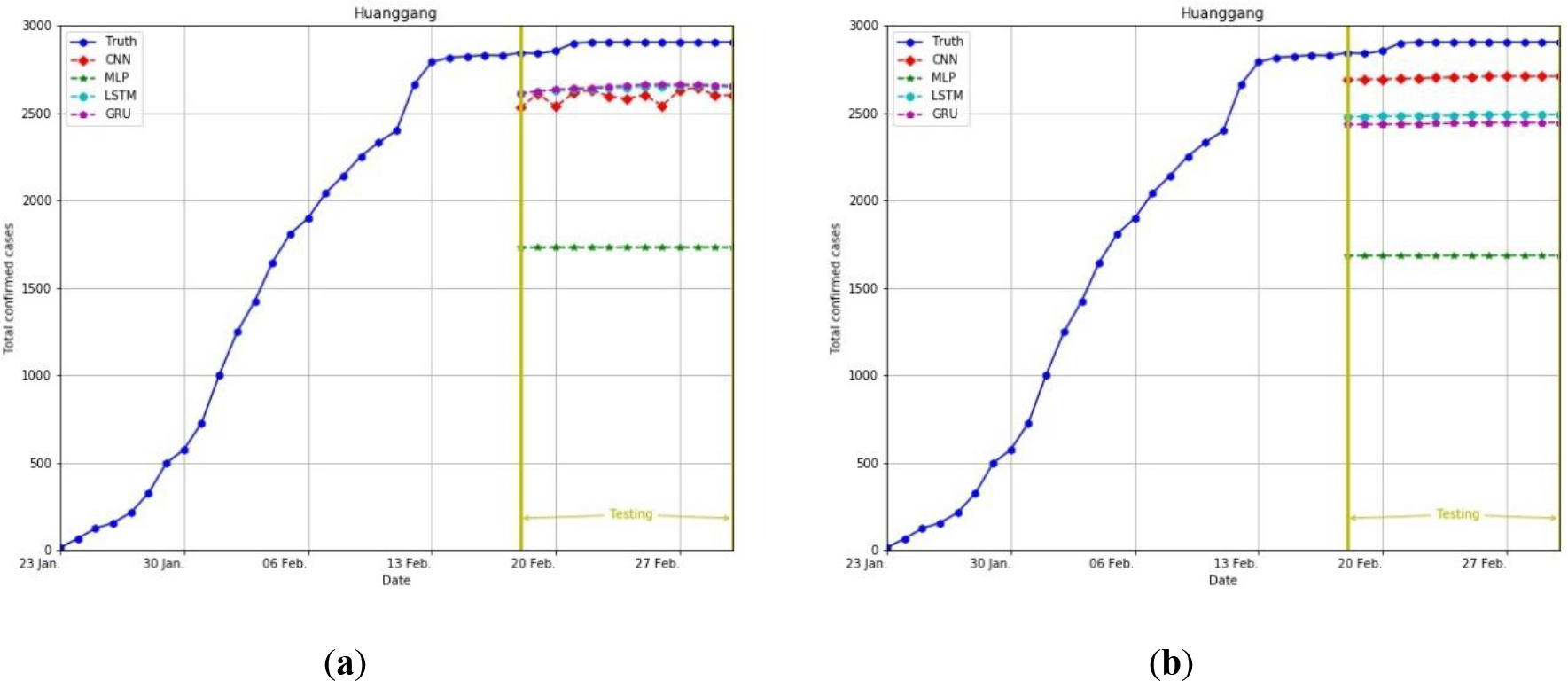
Comparison of prediction results for the cumulative number of confirmed cases in Huanggang City, Hubei Province, China. The comparison used the deep neural networks CNN, MLP, LSTM, and GRU. (a) Six important factors were adopted as the input layer: new confirmed cases, new deceased cases, new cured cases, cumulative confirmed cases, cumulative deceased cases, and cumulative cured cases. (b) Only one important factor was adopted as the input layer: cumulative confirmed cases.

**Fig. 8.**
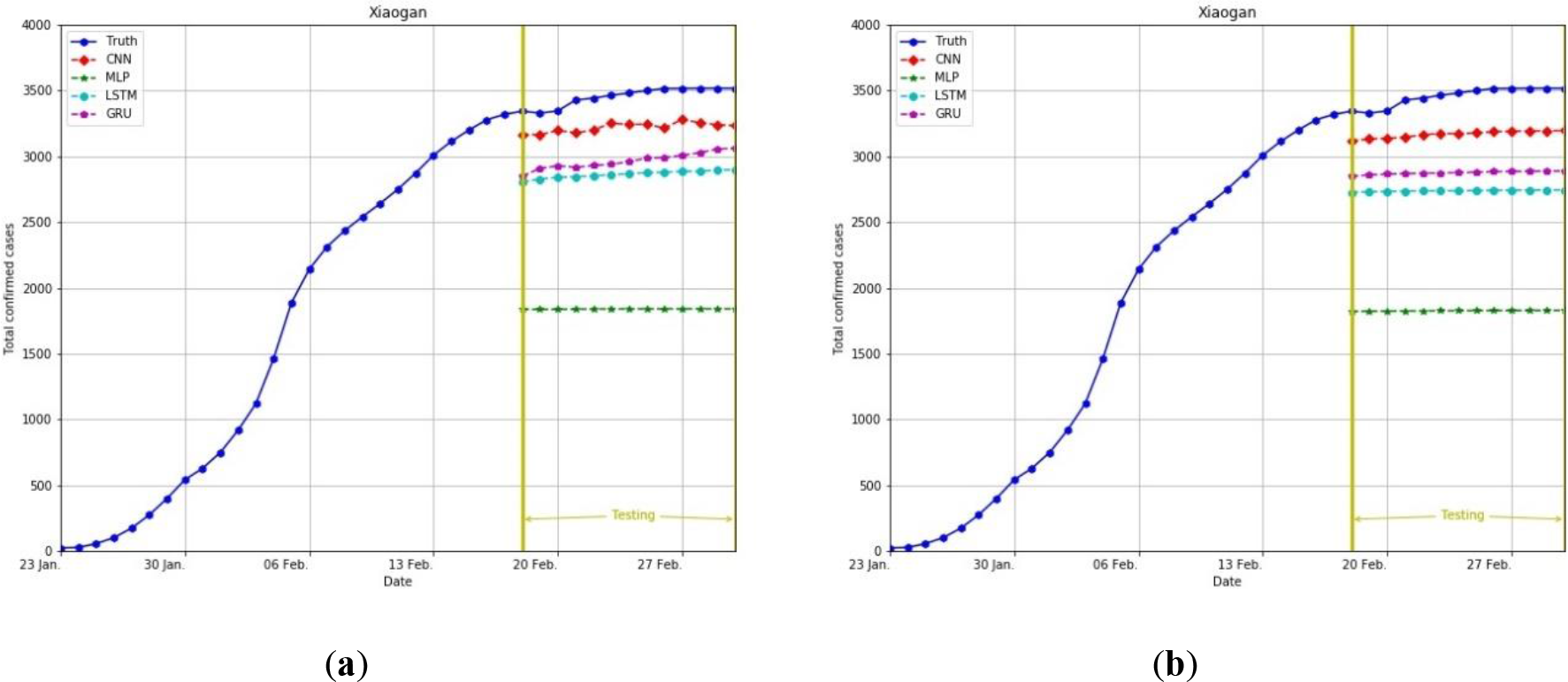
Comparison of prediction results for the cumulative number of confirmed cases in Xiaogan City, Hubei Province, China. The comparison used the deep neural networks CNN, MLP, LSTM, and GRU. (a) Six important factors were adopted as the input layer: new confirmed cases, new deceased cases, new cured cases, cumulative confirmed cases, cumulative deceased cases, and cumulative cured cases. (b) Only one important factor was adopted as the input layer: cumulative confirmed cases.

**Fig. 9.**
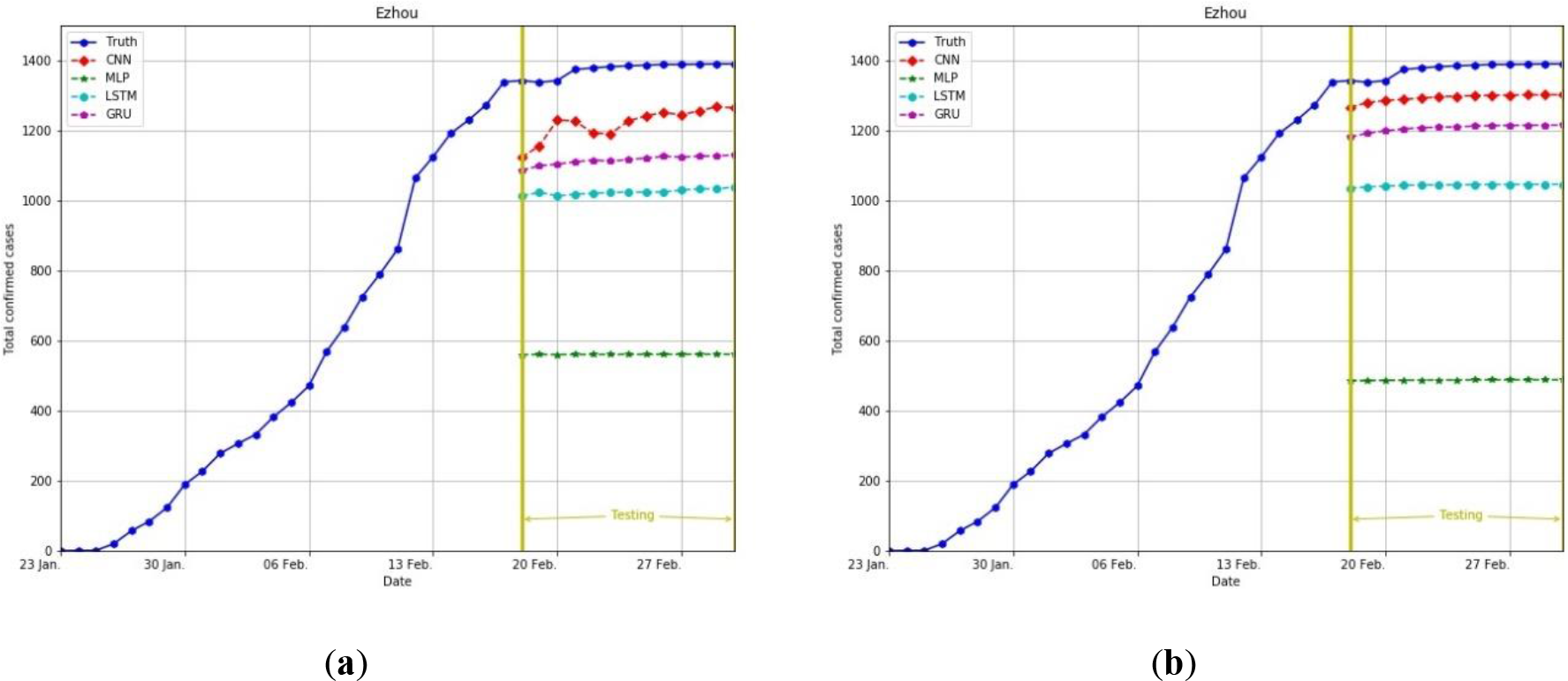
Comparison of prediction results for the cumulative number of confirmed cases in Ezhou City, Hubei Province, China. The comparison used the deep neural networks CNN, MLP, LSTM, and GRU. (a) Six important factors were adopted as the input layer: new confirmed cases, new deceased cases, new cured cases, cumulative confirmed cases, cumulative deceased cases, and cumulative cured cases. (b) Only one important factor was adopted as the input layer: cumulative confirmed cases.

**Fig. 10.**
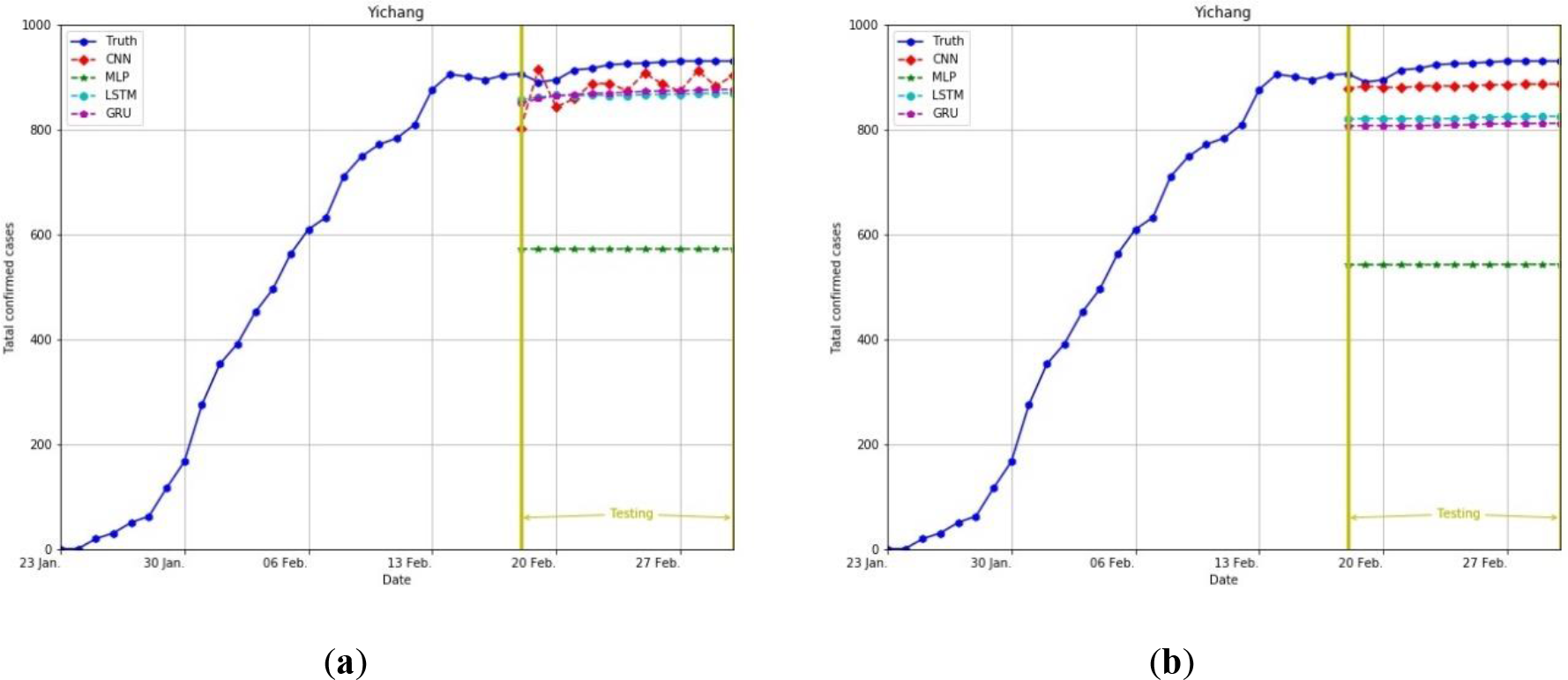
Comparison of prediction results for the cumulative number of confirmed cases in Yichang City, Hubei Province, China. The comparison used the deep neural networks CNN, MLP, LSTM, and GRU. (a) Six important factors were adopted as the input layer: new confirmed cases, new deceased cases, new cured cases, cumulative confirmed cases, cumulative deceased cases, and cumulative cured cases. (b) Only one important factor was adopted as the input layer: cumulative confirmed cases.

**Fig. 11.**
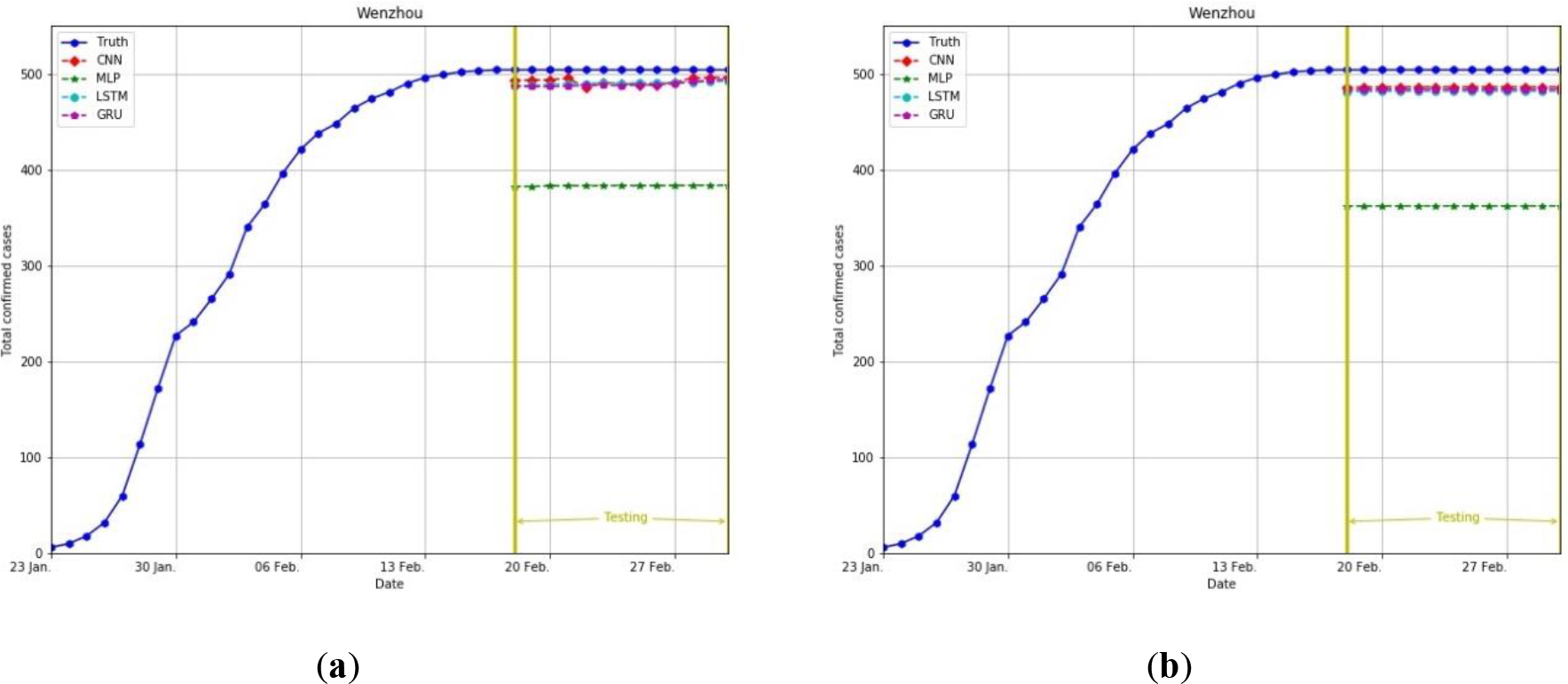
Comparison of prediction results for the cumulative number of confirmed cases in Wenzhou City, Zhejiang Province, China. The comparison used the deep neural networks CNN, MLP, LSTM, and GRU. (a) Six important factors were adopted as the input layer: new confirmed cases, new deceased cases, new cured cases, cumulative confirmed cases, cumulative deceased cases, and cumulative cured cases. (b) Only one important factor was adopted as the input layer: cumulative confirmed cases.

**Fig. 12.**
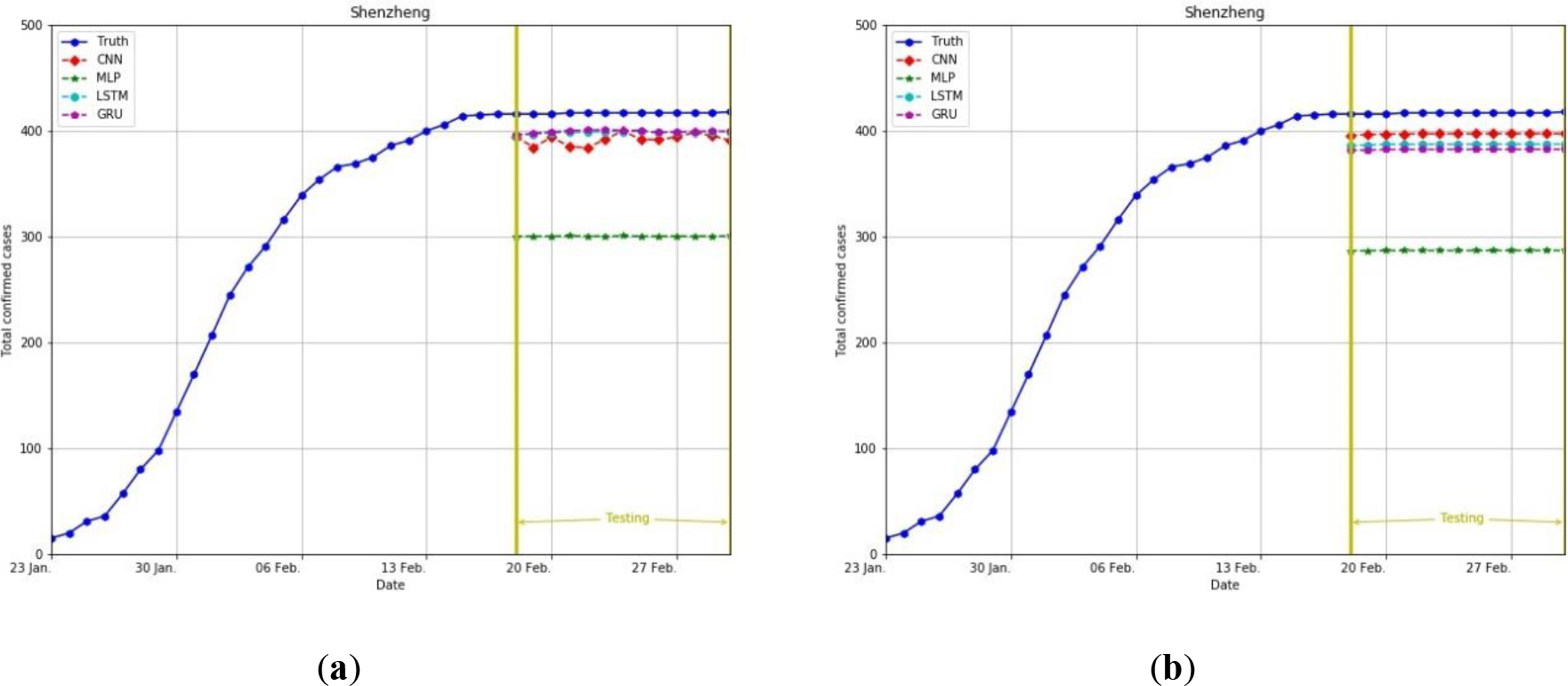
Comparison of prediction results for the cumulative number of confirmed cases in Shenzhen City, Guangdong Province, China. The comparison used the deep neural networks CNN, MLP, LSTM, and GRU. (a) Six important factors were adopted as the input layer: new confirmed cases, new deceased cases, new cured cases, cumulative confirmed cases, cumulative deceased cases, and cumulative cured cases. (b) Only one important factor was adopted as the input layer: cumulative confirmed cases.

Regarding the deep neural network prediction results for six and one-input factors, the MAE and RMSE values are listed in Tables 1 to 4. The MAE results from low to high are as follows: CNN (102.943, 284.573), GRU (2599.150, 1916.904), LSTM (2992.976, 3324.591), and MLP (5710.293, 5811.429). The RMSE results from low to high are as follows: CNN (109.439, 325.857), GRU (2602.666, 1924.361), LSTM (2994.851, 3331.925), and MLP (5715.125, 5816.079). The experiment results demonstrated that GRU and LSTM have decent efficacies and that CNN had the best performance. This experiment also demonstrated that the procedure of initial characteristic extraction through CNN and the subsequent input of characteristic values to the CNN structure greatly aid the prediction of the cumulative number of confirmed cases of COVID-19.

Fig. 13 (a) and (b) presents the radar charts of MAE values for the comparison between the output results and real values for six and one factors, respectively. Regardless of whether one or six factors were used, the prediction results, in terms of MAE, of the proposed CNN model were superior to those of its counterparts. For the proposed CNN model, the use of six factors yielded better prediction results, in terms of MAE, in comparison to the use of only one factor, particularly for Wuhan City.

**Fig. 13.**
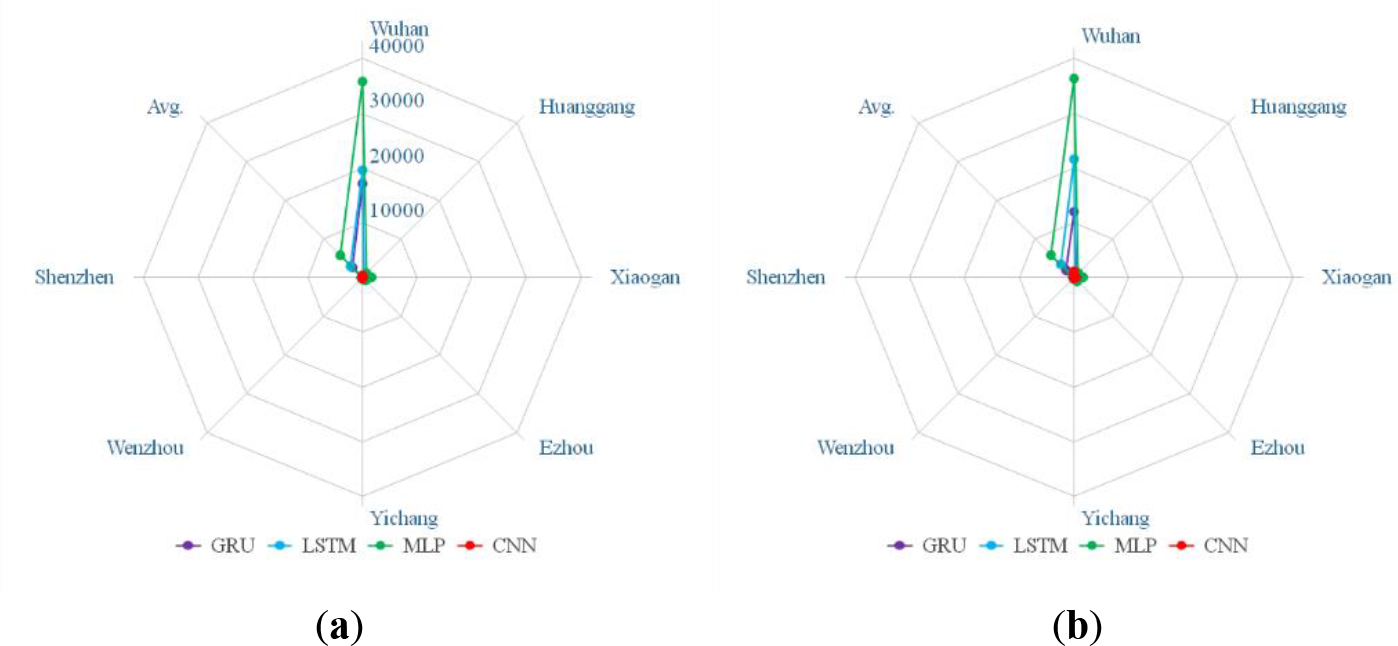
MAE radar charts. (a) Six factors were adopted as the input layer. (b) Only one factor was adopted as the input layer.

Fig. 14 (a) and (b) presents the radar charts of RMSE values for the comparison between the output results and real values for six and one factors, respectively. The results further demonstrated the excellent predictive performance of the proposed CNN model for the cumulative number of confirmed cases of COVID-19, making the model a valuable reference for other countries in their establishment of country-specific COVID-19 prediction models. In general, the prediction performances of the models from most favorable to least favorable were those for CNN, GRU, LSTM, and MLP.

**Fig. 14.**
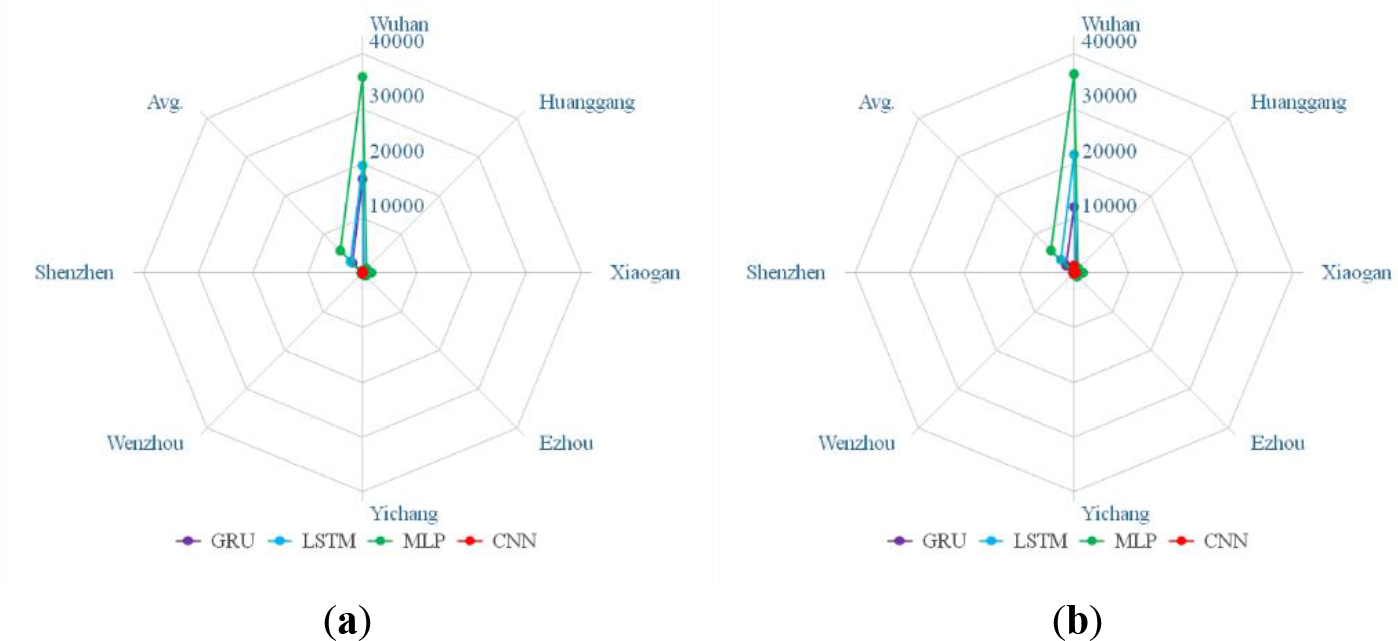
RMSE radar charts. (a) Six pieces of important information were adopted as the input layer. (b) Only one piece of information was adopted as the input layer.

## 5. Conclusions

A novel and multi-input CNN deep neural network model was proposed in this study to predict the cumulative number of confirmed cases of COVID-19. The cumulative number of confirmed cases in the next day is predicted according to the previous five days’ total number of confirmed cases, total confirmed new cases, total cured cases, total cured new cases, total deaths, and total new deaths. In the experiment, the datasets of those seven Chinese cities with severe confirmed cases—from Hubei Province, Guangdong Province, and Zhejiang Province—were used for the models’ training and prediction. Because the COVID-19 epidemic is ongoing, the algorithm proposed in this study can rapidly use small datasets to establish models with high predictive accuracy, different from many other studies. In addition, with the algorithm, a deep learning network prediction model for the number of confirmed cases of COVID-19 was established, and the verification and comparison were conducted among different deep learning algorithms. The accuracy and reliability of the deep learning algorithm were verified by having it predict the future trend of COVID-19. Furthermore, experiments for multiple cities with more severe confirmed cases in China indicated that the prediction model of this study had the lowest error rate among its counterparts tested. However, at the time of writing, countries other than China have had COVID-19 outbreaks, such as Italy, South Korea, Iran, Spain, France, and Germany. In the future, the establishment of more complete databases will lead to improvements to the proposed prediction model. For example, deep learning networks with a mixed structure can be introduced to establish more accurate models, which can be applied to more countries. The predicted trends can aid the containment of the COVID-19 epidemic and extend the scope of application of artificial intelligence.

## Data Availability

https://ncov.dxy.cn/ncovh5/view/pneumonia

